# Assessing the Real-World Performance of Xylazine Test Strips for Community-Based Drug Checking in Los Angeles

**DOI:** 10.1101/2025.06.28.25330471

**Authors:** Caitlin A. Molina, Joseph Friedman, Adam J. Koncsol, Ruby Romero, Morgan E. Godvin, Leslie Nuñez, Karmen G. Pang, Talya Tasini, Ezinne Okonkwo, Matthew Vu, Joshua Smith, Chelsea L. Shover

**Affiliations:** Department of General Internal Medicine and Health Services Research, University of California, Los Angeles; Department of Psychiatry, University of California, San Diego; University of California, Los Angeles

**Keywords:** Xylazine Test Strips, Concordance, Validation

## Abstract

**Background:** The veterinary sedative xylazine is increasingly found in illicit fentanyl and has been associated with numerous health harms. Xylazine test strips (XTS) are an emerging technology that can theoretically assist consumers in avoiding xylazine, but they require real-world validation. We leverage community-based drug checking program data to compare real-world XTS performance to ‘gold standard’ methods.

**Methods:** Samples were initially assessed by dissolving 1mg of drug product in 1mL water and dipping an XTS (“first generation” Wisebatch™) in the sample. Subsequently, confirmatory testing was performed by sending samples to the National Institute of Standards and Technology for qualitative analysis using direct analysis in real-time mass spectrometry (DART-MS). A subset was analyzed quantitatively with liquid chromatography gas spectrometry (LC-MS) to quantify xylazine, fentanyl, and other compounds.

**Results:** A total of n=1,570 drug samples were analyzed between June 2023 and May 2025, and a total of n=801 XTS were used. N=715 comparisons between xylazine test strips and mass spectrometry results could be made, including n=333 among samples that tested positive for fentanyl. Of these, n=63 samples were confirmed to contain xylazine by mass spectrometry, of which the majority contained low concentrations (average concentration 2.3%; 78% of samples contained less than <1% xylazine by weight). Of the 63, n=34 were correctly identified as positive by XTS, yielding sensitivity of 54.0%. Of n=270 xylazine negative samples, n=235 were correctly categorized (specificity=87.0%). Most false positives occurred with lidocaine present.

**Conclusions:** In our sample, with a large percentage of low concentration xylazine samples, “first generation” Wisebatch XTS had a relatively low sensitivity, but higher specificity. This highlights the value of confirmatory testing and the complicated and often confusing nature of point-of-care test strips for novel substance detection. Lot testing and validation studies are needed to improve quality control in this area.

## Introduction

Xylazine is an alpha-2 adrenergic agonist that causes fast-acting sedation, for which it is used widely as a veterinary sedative[1]. Xylazine is not approved for use in humans by the US Food and Drug Administration. The drug has increasingly been found alongside fentanyl in illicit drug samples and overdose toxicology[2–4]. The addition of xylazine to fentanyl was initially more common in Philadelphia before spreading to other parts of the Eastern US. However, xylazine-adulterated fentanyl has now spread to nearly every state in the US[5], including the West Coast of the US and Northern Mexico[4,6,7].

Xylazine has been associated with numerous health harms for people who use drugs, such as more complicated overdose management[6,8,9] soft tissue infections[10–12], and complex withdrawal syndromes[13]. Furthermore, the subjective experience of xylazine with fentanyl is qualitatively different from that of using opioids alone. For these reasons, many people who use drugs wish to avoid xylazine, but are often unable to do so due to a lack of knowledge of the substance, inability to identify samples that contain it, or a lack of options in the illicit drug market that do not contain it[2,14].

Xylazine test strips (XTS) are an emerging technology that can theoretically assist consumers in avoiding xylazine. They are produced by several manufacturers and are low-cost, usually about $2 USD per strip. They leverage immunoassay technology to offer a presence/absence binary result in less than one minute, describing if xylazine is present in a drug sample[15–17].

Nevertheless, these strips require validation to assess their utility in real-world settings, as evidence regarding their effectiveness has been mixed.

A small pilot study in Rhode Island of n=41 drug samples found xylazine present in n=18 samples according to a ‘gold standard’ approach leveraging mass spectrometry, however XTS (the “first generation” of strips produced by BTNX) were only reliably able to detect xylazine in the n=4 samples where it was a ‘major component’ (>30% by weight) of the drug sample[15]. Despite the small sample size, that study suggested that XTS may not be able to detect xylazine when it is found at lower concentrations in drug samples[15]. A pilot sample from Tijuana, Mexico, compared two brands of XTS to direct analysis in real-time mass spectrometry (DART-MS) [7]. In that sample, n=12 samples tested positive for xylazine on DART-MS confirmatory testing. Of these, Wisebatch XTS correctly identified all n=12, however, also showed n=3 false positives when lidocaine was present. SafeLife XTS had n=0 false positives, but did have n=1 false negative (identifying n=11 total xylazine positive samples). A report from British Columbia found that among n= 152 samples tested using first-generation BTNX XTS and confirmatory testing, sensitivity was 97.7% and specificity was 66.7%. That study also found that a relatively high rate of false positives were likely caused by the presence of ortho-methylfentanyl[18]. The fairly heterogeneous findings noted above highlight that further data are needed to understand the real-world performance of XTS in distinct illicit drug markets.

Here we provide the first US West Coast-based study of the real-world performance of XTS on the illicit drug market, leveraging data from a community-based drug checking program, Drug Checking Los Angeles. This study draws on n=715 samples with XTS and confirmatory results—the largest sample to date, to our knowledge.

## Methods

Drug samples were provided confidentially by anonymous clients using the services of a community-based drug checking program based at several sites in Los Angeles, California. Test strips for several substances, including xylazine, were employed on-site. XTS were initially used for all samples, and later limited to only those samples bought as or otherwise expected to be fentanyl. Samples were initially assessed by dissolving 1mg of drug product in 1 mL of water and dipping an XTS (from the “first generation” of Wisebatch™ brand strips) in the sample. The strips were read by trained technicians, and a second strip was used in the rare case of an indeterminate result (e.g., faint test line, technician disagreement, etc.). No invalid results were observed during the study. The xylazine test strips evaluated were purchased from Wisebatch Harm Reduction (Costa Mesa, CA), sold as having a cut-off value of 1000ng/mL. All strips were from lot D2306292, with an expiration date listed as 2025-06-25. None of the strips evaluated were expired at the time of use.

Confirmatory testing was pursued for the majority of samples (unless clients declined this service), wherein swabs/vials taken from drug samples were sent to the National Institute of Standards and Technology (NIST) for laboratory-based qualitative testing using DART-MS. A subset also underwent quantitative analysis with liquid-chromatography mass spectrometry (LC-MS) [only those samples for which sufficient volume was available, approximately 5mg].

The confirmatory testing methodologies employed here have been previously described [19,20]. Briefly, the spectra produced by DART-MS analysis were compared against libraries of over 1,300 substances, including pharmaceutical and illicit drugs, adulterants, cutting and bulking agents, precursor chemicals, and other substances (e.g., adhesives, food products, etc.).

An LC-MS quantification panel included xylazine, fentanyl, fentanyl precursor chemicals, other illicit drugs, and common adulterants. Substances found to be present, but below the limit of quantification were imputed at 0.1% prevalence. The limit of detection, and limit of quantification of xylazine in the LC-MS approach used here, were 0.01% by mass and 0.1% by mass, respectively[21]. The LOD of DART-MS for the detection of xylazine was 1ng/mL[21].

Data analysis, including descriptive statistics and graphing, were conducted using R 4.1.1. The UCLA Institutional Review Board reviewed and approved this project (protocol IRB-22-0760) and additionally determined that aspects of this work constituted public health surveillance and not human subjects research.

## Results

A total of n=1,570 samples were analyzed between June 2023 and May 2025, of which n=545 were fentanyl-positive on DART-MS. A total of n=801 XTS were used, and of these n=715 had confirmatory DART-MS data available for the samples (including n=333 fentanyl-positive samples per DART-MS). A total of N=119 samples were xylazine-positive on DART-MS (about 20% of fentanyl-positive samples). N=87 samples had xylazine quantification data available by mass spectrometry. The majority contained low concentrations of xylazine. The average xylazine concentration was 2.3% by mass (range: below LOQ [≈0.1%]-56.9%). N=68 (78%) of samples contained less than <1% xylazine by weight.

Among the n=333 XTS testing results available used for the primary analysis, given that they were on fentanyl-positive samples per DART-MS, 100% also had xylazine testing results on DART-MS available. Of these, n=63 samples were confirmed to contain xylazine by mass spectrometry. Among this sample, n=34 were correctly identified as positive by XTS, yielding sensitivity of 54.0%. Of n=270 xylazine negative samples, n=235 were correctly categorized (specificity=87.0%). Positive predictive value was 49.3% and negative predictive value was 89.0%. Performance statistics on the full sample of n=715 XTS with DART-MS results available were similar (see supplemental table).

False negative results on XTS qualitatively appeared to be more common among samples with <5% xylazine concentration and were less common with samples above 5% concentration (Figure 1). However small numbers of samples with higher xylazine concentrations precluded formal hypothesis testing of these differences. Among n=36 false positive results (in the full sample; see supplement), n=33 samples (91.7%) were found to contain lidocaine on DART-MS (supplemental table 2). These false positives were seen across a broad range of lidocaine concentrations (supplemental figure 1).

**Figure 1.**
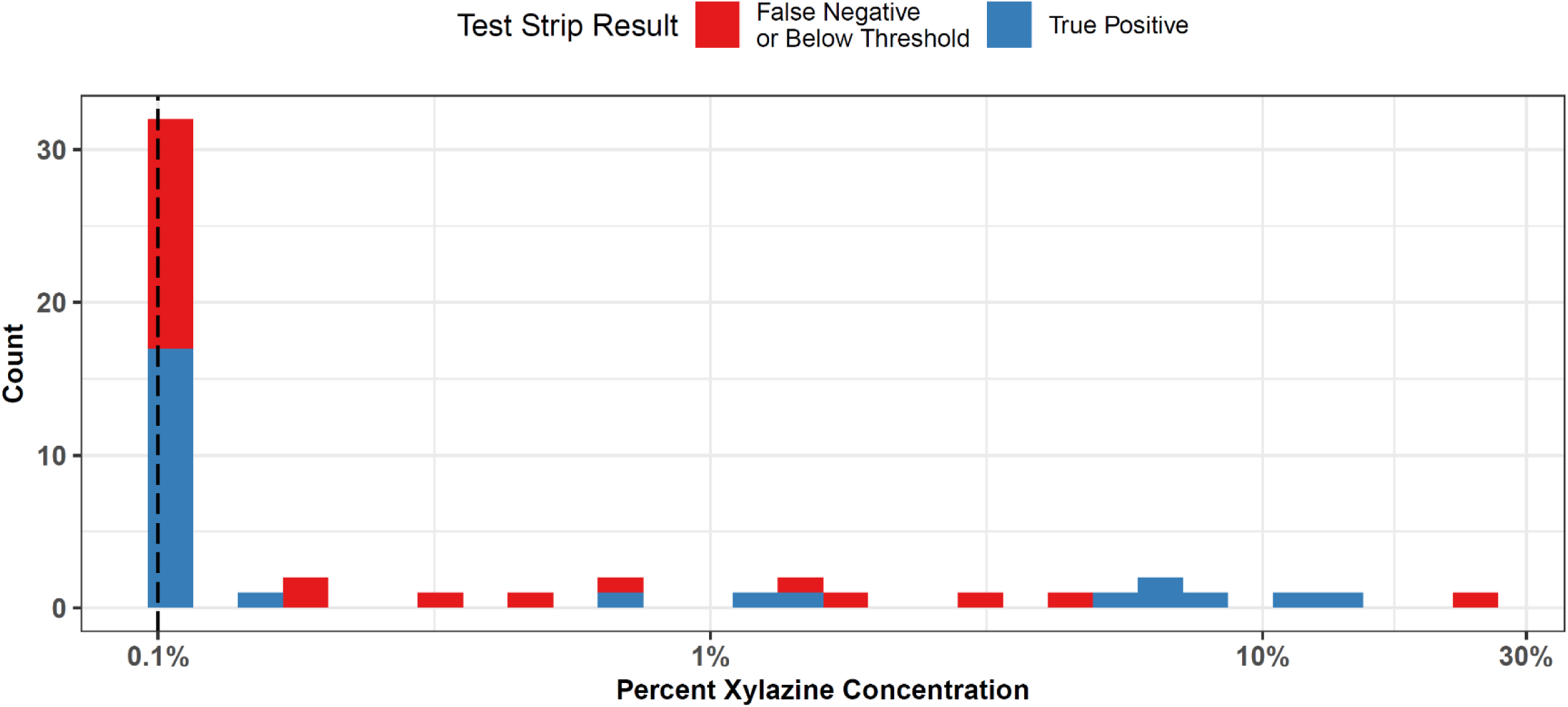
**Xylazine Test Strip Results by Concentration** The distribution of xylazine concentration (by weight) according to LC-MS is shown among samples with xylazine test strip results available. A log scale is used on the x-axis to show the percent concentration by mass of xylazine. Results are shown separate by false negative (shown in red) and true positives (shown in blue). Of note, some ‘false negatives’ may be appropriate as the concentration of xylazine is below the stated LOD of the test strips. The stated LOD of the XTS of 1000ng/mL, which given our preparation of 1mg of sample per 1mL of water, would correspond to 0.1% xylazine by mass, which is shown with a dashed vertical line.

## Discussion

This study adds to a small but growing literature describing the real-world performance of XTS, compared to confirmatory testing using mass spectrometry[7,15,18]. With n=333 comparison points in the primary sample (and n=715 comparison points in the supplementary analysis not limited to fentanyl-positive samples), this study represents the largest to-date on this topic to our knowledge, and the first on the West Coast of the U.S. In our sample, with a large percentage of low concentration xylazine samples, “first generation” Wisebatch XTS had a relatively low sensitivity, but high specificity. This aligns with a previous smaller study from Rhode Island, which found that XTS may be prone to a high rate of false negative readings among samples that had lower concentrations of xylazine[15]. Our results stand in contrast to another recent report from British Canada, which found a high sensitivity, but low specificity[18]. Nevertheless, the percentage xylazine concentration among samples was not reported in that study, therefore those results may reflect only higher-concentration samples. It is also plausible that each study tested different batches/lots of XTS, also affecting XTS performance assessment, as lot-to-lot variability of immunoassay test strip sensitivity has been documented[22]]. In response to early results showing false positives in response to certain cutting agents, such as lidocaine, some XTS manufacturers created a “second generation” of strips designed to reduce such false positives. Further study of subsequent generations of XTS is needed. Repeating similar studies to the current analysis for subsequent generations of test strips would be valuable. If they continue to show that current generations of XTS are unlikely to detect low concentrations of xylazine, there may be value in the development of XTS with lower detection thresholds. Consistent with other studies of “first generation” XTS we find that lidocaine was present in the vast majority of false positive XTS results[7]. We found lidocaine present at a wide range of concentrations for false positive XTS results, without a clear threshold effect.

This study is limited by the nature of the illicit drug market in Los Angeles, which appears to have an abundance of fentanyl samples with low xylazine concentration. This limited our ability to assess XTS performance at higher concentrations. Given little data nationally on quantitative testing of xylazine in other drug markets, it remains to be seen how similar the illicit drug market is to other environments, and such differences may affect XTS performance. The study is also only representative of the specific clients who participate in services at Drug Checking Los Angeles sites and may not reflect the entirety of the Los Angeles or West Coast illicit drug market. The clinical relevance of frequent, low-concentration xylazine consumption is also not known, given limited study of xylazine among humans.

Given the low concentration of xylazine in the Los Angeles fentanyl supply, xylazine test strips with lower detection thresholds may be useful. The stated detection threshold for the Wisebatch strips leveraged in this study was 1000 ng/mL. Therefore, given our use of 1mg/mL of water, they should theoretically detect the presence of xylazine when it was at least 0.1% by mass of the sample. The DART-MS approach we employed is considerably more sensitive, with a xylazine detection threshold of 1 ng/mL[21]. Therefore, samples with between 1 ng/mL and 1000 ng/mL would likely be positive on DART-MS and negative on the xylazine test strips we used here. Given that our LOQ for LC-MS was 0.1% by mass, and not all samples with DART-MS results had LC-MS quantification available, we are not able to characterize the percent by mass of these extremely low concentration samples. This represents a limitation to our study. For this analysis, we have labeled these discordant cases ‘false negatives’, nevertheless, this designation depends on if low concentration xylazine is of importance and should be detected. If individuals consuming xylazine care about low concentration xylazine, then the strips should have a lower detection threshold and these might be considered ‘false negatives’. In that case, our results would suggest that for the measurement of the low concentration xylazine found in fentanyl, using a higher concentration solution, e.g. 2 mg drug product/mL may also be warranted, if more sensitive strips are not available. If individuals consuming substances only care about higher concentrations of xylazine, then these might be considered appropriate negatives, and the detection threshold may be appropriate at its current level. This will depend on the currently unknown clinical importance of low-dose xylazine exposure, which future studies will be needed to elucidate. Nevertheless, we did observe many examples of xylazine concentration above 0.1% by mass which were false negatives on xylazine test strips, as well as true positives below the stated LOD of the XTS, as well as false positives, therefore still indicating overall relatively poor discriminatory ability of the XTS tested in this study.

The poor performance of test strips in this context highlights the value of laboratory-based testing, especially as subsequent generations of XTS are still in development. The landscape of point-of-care test strips for novel substance detection remains complicated and often confusing for end consumers. Nevertheless, there is clear interest in and demand for rapid technologies that can detect new harmful substances, such as XTS[14]. Follow-up studies on subsequently released test strip products, in more diverse xylazine markets, are needed. Furthermore, in response to the mixed and confusing landscape of test strip technologies, other authors have argued that lot testing and validation studies should be undertaken by the harm reduction community[23–25].

An additional consideration for the widespread implementation of XTS along with other test strips for novel substances lies in the complexities around accurate field measurements and dilution strategies. Although in this study, efforts were taken to ensure that 1mg of product and 1mL of solution were used (see supplemental methods), it is possible that some variation in the quantity of sample was present due to operator imprecision. Furthermore, in many field-based contexts, where precise measurement tools are not available, we would expect this variation to increase considerably. Additionally, many individuals prepare one drug sample for concurrent testing with various test strips, (e.g. xylazine, fentanyl, nitazenes) and the use of various dilutions may be impractical. This is both a potential source of variation between studies in literature (which have distinct preparation protocols), and a key source of potential error for real-world scenarios. A pragmatic approach may be found in the field of harm reduction setting standard dilutions (e.g. 1mg/mL), the widespread distribution of scoops and pipettes that facilitate these measurement, and broader testing protocols oriented towards these specific dilutions. Additionally, testing procedures should be developed and implemented keeping in mind that in real-world conditions, precise quantities of drug sample may be difficult to guarantee. Therefore, testing materials should be robust to a range of testing quantities around the indicated amount.

By highlighting arguably poor performance of some XTS strips in a specific context, our study adds additional evidence that standardization, validation studies, and guidance for service providers seeking to use test strips in harm reduction contexts are needed.

**Table 1.**
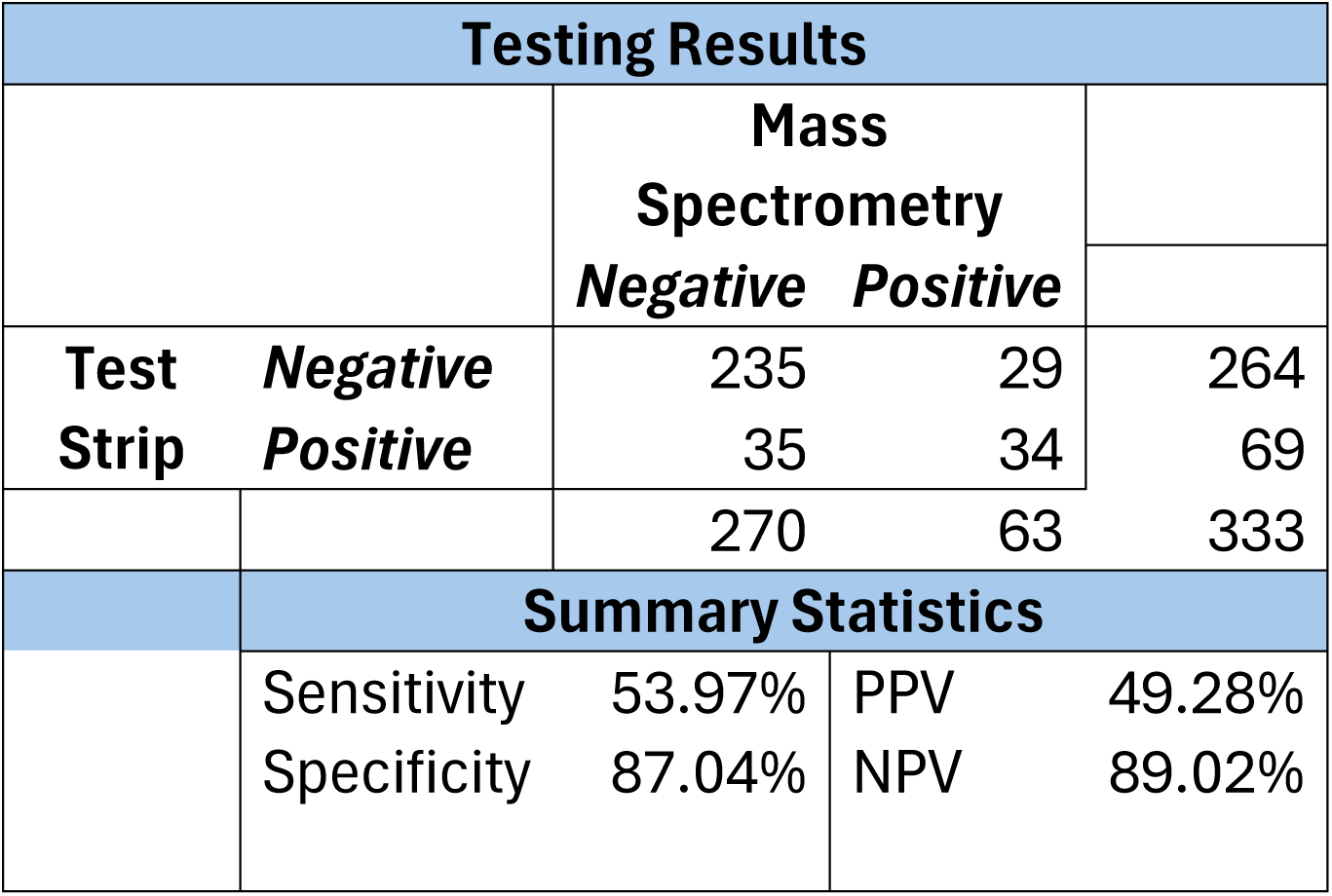
Predictive Performance of Xylazine Test Strips (Among Fentanyl-Positive Samples) A comparison is shown between results from xylazine test strips to confirmatory testing based on DART-MS. Sensitivity, specificity, positive predictive value, and negative predictive value are also calculated. Row and column marginal sums are also shown.

| Testing Results |  |  |  |  |
| --- | --- | --- | --- | --- |
|  |  | Mass Spectrometry |  |  |
|  |  | Negative | Positive |  |
| Test Strip | Negative | 235 | 29 | 264 |
|  | Positive | 35 | 34 | 69 |
|  |  | 270 | 63 | 333 |
| Summary Statistics |  |  |  |  |
|  | Sensitivity | 53.97% | PPV | 49.28% |
|  | Specificity | 87.04% | NPV | 89.02% |

## Data Availability

The underlying data are sensitive and cannot be shared. However, summary statistics may be provided by the authors upon reasonable request.

## Declarations

### Ethics approval and consent to participate

The UCLA Institutional Review Board reviewed and approved this project (protocol IRB-22-0760) and additionally determined that aspects of this work constituted public health surveillance and not human subjects research.

### Availability of data and materials

Data are sensitive and cannot be shared. However, researchers may request summary data from the authors to be provided upon reasonable request.

## Funding

JRF received funding from the National Institute on Drug Abuse (DA049644; 1U01DA063078) and the National institute of Mental Health (MH101072). AJK received educational support through the NIH/National Center for Advancing Translational Science (NCATS) UCLA CTSI (TL1TR001883). CLS received support from the National Institute on Drug Abuse (K01DA050771 and R01DA057630). This work was supported by the Centers for Disease Control and Prevention as part of Overdose Data to Action: LOCAL (CDC-RFA-CE-23-0003), and made possible through an equipment grant from the James B. Pendleton Charitable Trust to the UCLA AIDS Institute and UCLA Center for AIDS Research.

### Authors’ contributions

CLS conceptualized the article, obtained grant funding, and provided overall supervision. All authors participated in the data collection. CAM and JRF wrote the initial draft. CAM and JRF conducted the statistical analysis. CAM, RR, and AJK handled data management. All authors critically reviewed and approved the manuscript.

## Acknowledgements

We thank the staff, volunteers, and community partners of Drug Checking Los Angeles for their work to provide services for this client population, and the clients for their engagement in services and efforts to improve them moving forward. We gratefully acknowledge the Rapid Drug Analysis and Research (RADaR) program of the National Institute of Standards and Technology for performing all laboratory testing discussed in this analysis.

## Supplemental Methods

### Test Strip Procedure

Samples were prepped in the field using a 5-10 mg microscoop to place 1 mg of sample into a 1.5 mL graduated Eppendorf vial. Drug checking technicians were trained to identify what 1 mg of sample in the tube would look like, using an image of an Eppendorf tube with 1 mg of sugar weighed on a scale. After 1 mg of sample were added to the Eppendorf vial, 1 mL of sterile water was added from a disposable water ampoule, using the measurements on the side of the vial. The vial was capped, and technicians manually agitated the vial for 10 seconds to ensure the contents of the vial were adequately mixed. A single test strip was dipped into the vial, inserting the wavy liquid icon below the max line of the test strip until the technician saw water begin to run up the test strip, about 15 seconds. Once water ran up the strip, the test strip was removed from the vial and placed on a flat, nonabsorbent surface. After 2 minutes, test strip results were read. Senior technicians oversaw all drug checking procedures, to ensure that the appropriate procedures were followed.

### Steps from training manual for XTS strips

1. 1 mg of the same sample and 1.0 ml of filtered water were added to an Eppendorf vial. The vial was agitated manually for ten seconds.
2. We then employed xylazine test strips from Wisebatch to check each sample for the presence of xylazine. The wavy side of each strip was inserted into the dissolved solution until water began to run up the strip, about 15 seconds. Technicians are trained to dip the strip taking care to not insert it past the thick blue max line. After water began to run up the XTS, the strip was placed flat on a clean, non-absorbent surface to allow the solution to run up the strip to run via capillary action.
3. After 2 minutes, two trained technicians read the test strip results. In the rare case of disagreement between technicians, or an inconclusive or invalid result, a second strip was employed, providing a definitive result in all cases.
4. Interpretation of test strip results: One line is positive, two lines are negative, and the test is invalid if no lines show or there is no control line.

### Sample Disposal Procedure

To dispose of the samples, we used nontoxic DisposeRx powder to render the drug samples inert in a viscous gel.

1. The sample powder, sample solution in the Eppendorf vial, a DisposeRx packet, and 15 mL of filtered water were added to a 22 mL vial at the end of each drug checking clinic.
2. The vial was then agitated manually until the gel formed.
3. This vial was then safely discarded into the trash.

### Laboratory Sample Preparation

1. To collect samples for lab-based testing, drug checking technicians used a 5mg microscoop to place 1 scoop of sample in a labelled 2mL amber glass vial containing about 0.5mL of acetonitrile.
2. The vial is then tightly capped and sealed with a strip of parafilm to prevent leakage.
3. This vial is then placed in the corresponding labelled plastic envelope.
4. All sample envelopes are then placed in a clear zippered bag with a sorbent material, to be submitted to NIST for qualitative analysis with DART-MS and quantitative analysis with LC-MS/MS.

## Supplemental Results

**Supplemental Table 1.** Predictive Performance of Xylazine Test Strips Among Full Sample, Not limited to Fentanyl Positive Samples A comparison is shown between results from xylazine test strips to confirmatory testing based on DART-MS. Sensitivity, specificity, positive predictive value (PPV), and negative predictive value (NPV) are also calculated. Row and column marginal sums are also shown.

| Testing Results |  |  |  |  |
| --- | --- | --- | --- | --- |
|  |  | Mass Spectrometry |  |  |
|  |  | Negative | Positive |  |
| Test Strip | Negative | 614 | 31 | 645 |
|  | Positive | 36 | 34 | 70 |
|  |  | 650 | 65 | 715 |
| Summary Statistics |  |  |  |  |
|  | Sensitivity | 52.31% | PPV | 48.57% |
|  | Specificity | 94.46% | NPV | 95.19% |

**Supplemental Table 2.** Drug Contents for Samples with False Positive XTS Results For the n=36 samples with a positive XTS results and no xylazine found on DART-MS, the sample contests according to DART-MS are shown. Binary presence/absence indicators are shown for two substances known to cause false positives among XTS in the literature, namely lidocaine and ketamine. Lidocaine was found among n=33, representing 91.7% of false positives. Ketamine was found among n=1 sample.

| # | Drug Contents on DART-MS | Lidocaine | Ketamine |
| --- | --- | --- | --- |
| 1 | Cocaine, Lidocaine, Ketamine, Benzoylecgonine, Acetaminophen | 1 | 1 |
| 2 | 4-ANPP, Acetaminophen, Caffeine, Cocaine, Fentanyl, Mannitol, Norfentanyl, Theobromine, Lidocaine, Aniline | 1 | 0 |
| 3 | Acetaminophen, Cocaine, Fentanyl, Lidocaine, Quetiapine, Caffeine | 1 | 0 |
| 4 | 4-ANPP, Acetaminophen, Fentanyl, Lidocaine, Fluorofentanyl | 1 | 0 |
| 5 | 4-ANPP, Acetaminophen, Fentanyl, Lidocaine | 1 | 0 |
| 6 | 4-ANPP, Acetaminophen, Caffeine, Fentanyl, Lidocaine, Norfentanyl, Methamphetamine, Aniline | 1 | 0 |
| 7 | Acetaminophen, Fentanyl, Lidocaine, 4-Anpp, Phenethyl 4-Anpp | 1 | 0 |
| 8 | 4-ANPP, Acetaminophen, Bis(2,2,6,6-Tetramethyl-4-Piperidyl) Sebacate, Fentanyl, Lidocaine, Methamphetamine, Mannitol, Phenethyl 4-Anpp | 1 | 0 |
| 9 | 4-ANPP, Acetaminophen, Bis(2,2,6,6-Tetramethyl-4-Piperidyl) Sebacate, Fentanyl, Lidocaine, Mannitol, Aniline, Fluorofentanyl | 1 | 0 |
| 10 | 4-ANPP, Acetaminophen, Bis(2,2,6,6-Tetramethyl-4-Piperidyl) Sebacate, Fentanyl, Lidocaine, Mannitol, Aniline, Fluorofentanyl, Ethyl 4-Anpp | 1 | 0 |
| 11 | 4-ANPP, Acetaminophen, Fentanyl, Fluorofentanyl, Lidocaine | 1 | 0 |
| 12 | Fentanyl, Lidocaine, Mannitol, 4-ANPP, Phenethyl 4-ANPP | 1 | 0 |
| 13 | Acetaminophen, Fentanyl, Lidocaine | 1 | 0 |
| 14 | 4-ANPP, Bis(2,2,6,6-Tetramethyl-4-Piperidyl) Sebacate, Fentanyl, Lidocaine, Phenacetin | 1 | 0 |
| 15 | Bis(2,2,6,6-Tetramethyl-4-Piperidyl) Sebacate, Caffeine, Lidocaine, Mannitol, Fentanyl | 1 | 0 |
| 16 | 4-ANPP, Acetaminophen, Fentanyl, Lidocaine, Ethyl 4-ANPP | 1 | 0 |
| 17 | Acetaminophen, Fentanyl, Lidocaine, Mannitol, NPP | 1 | 0 |
| 18 | Acetaminophen, Fentanyl, Lidocaine, Fluorofentanyl, 4-ANPP | 1 | 0 |
| 19 | 4-ANPP, Bis(2,2,6,6-Tetramethyl-4-Piperidyl) Sebacate, Fentanyl, Lidocaine, 1-Phenethylpiperidin-4-ol, Mannitol | 1 | 0 |
| 20 | Acetaminophen, Bis(2,2,6,6-Tetramethyl-4-Piperidyl) Sebacate, Fentanyl, Lidocaine, 4-ANPP | 1 | 0 |
| 21 | Acetaminophen, Fentanyl, Lidocaine, 4-ANPP, NPP, Bis(2,2,6,6-Tetramethyl-4-Piperidyl) Sebacate, Aniline, Mannitol | 1 | 0 |
| 22 | 4-ANPP, Acetaminophen, Fentanyl, Lidocaine | 1 | 0 |
| 23 | 4-ANPP, Acetaminophen, Fentanyl, Lidocaine | 1 | 0 |
| 24 | 4-ANPP, Acetaminophen, Bis(2,2,6,6-Tetramethyl-4-Piperidyl) Sebacate, Fentanyl, Lidocaine, Mannitol, Norfentanyl, Fluorofentanyl | 1 | 0 |
| 25 | Fentanyl, Lidocaine | 1 | 0 |
| 26 | Fentanyl, Lidocaine, 4-ANPP; Aniline; Norfentanyl; Bis(2,2,6,6-Tetramethyl-4-Piperidyl) Sebacate | 1 | 0 |
| 27 | Acetaminophen, Fentanyl, Lidocaine, 4-ANPP | 1 | 0 |
| 28 | 4-ANPP, Acetaminophen, Fentanyl, Lidocaine, Bis(2,2,6,6-Tetramethyl-4-Piperidyl) Sebacate | 1 | 0 |
| 29 | Acetaminophen, Bis(2,2,6,6-Tetramethyl-4-Piperidyl) Sebacate, Fentanyl, Lidocaine, MDMA, Norfentanyl 4-ANPP | 1 | 0 |
| 30 | 4-ANPP, Acetaminophen, Fentanyl, Lidocaine, Mannitol, Norfentanyl | 1 | 0 |
| 31 | Acetaminophen, Bis(2,2,6,6-Tetramethyl-4-Piperidyl) Sebacate, Fentanyl, Lidocaine, MDMA, Norfentanyl, 4-ANPP | 1 | 0 |
| 32 | 4-ANPP, Fentanyl, Lidocaine | 1 | 0 |
| 33 | 4-ANPP, Fentanyl, Lidocaine | 1 | 0 |
| 34 | Fentanyl, Caffeine, 4-ANPP, Mannitol | 0 | 0 |
| 35 | 4-ANPP, Acetaminophen, Fentanyl, Fluorofentanyl | 0 | 0 |
| 36 | 4-ANPP, Acetaminophen, Fentanyl, Fluorofentanyl | 0 | 0 |

**Supplemental Figure 1.**
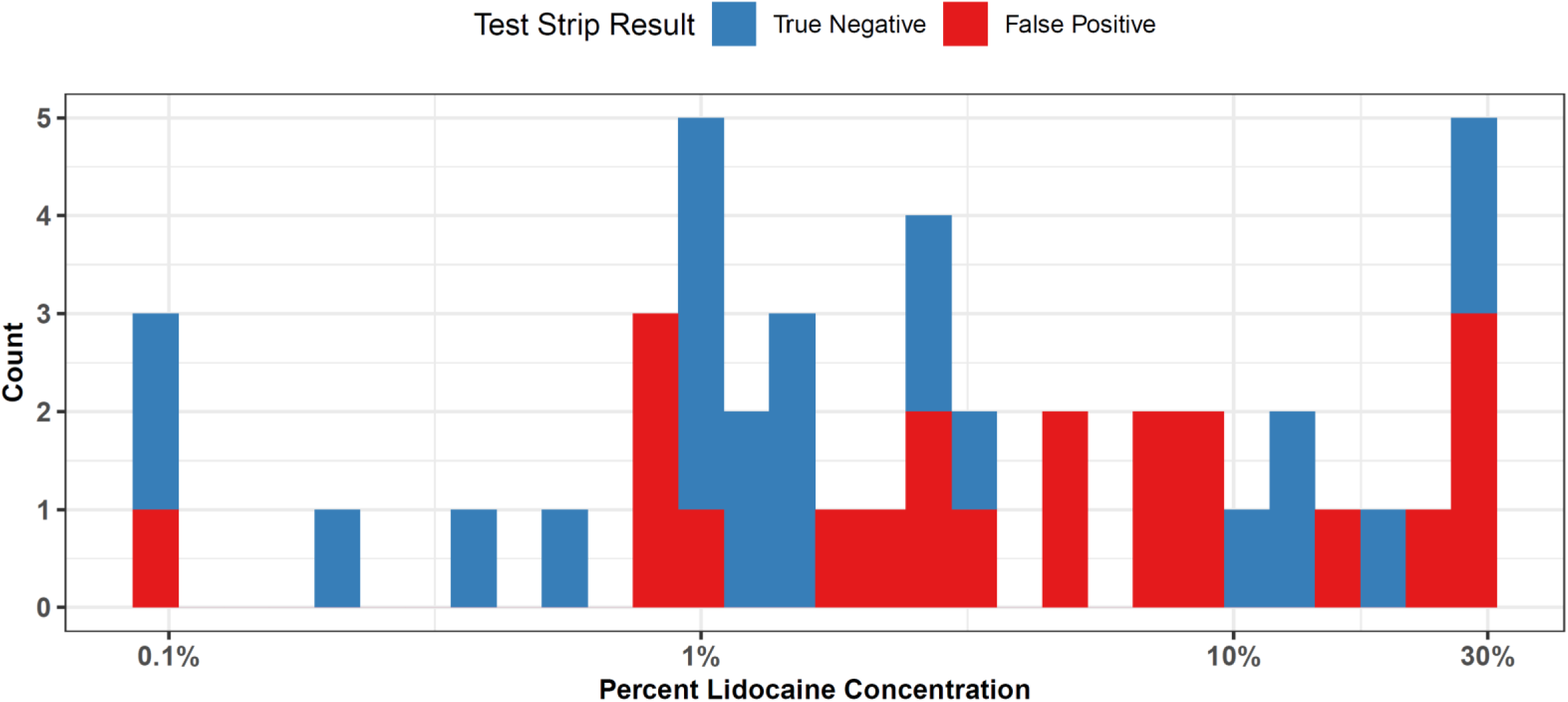
Xylazine Test Strip Results by Lidocaine Concentration (Among Xylazine Negative Samples) The distribution of lidocaine concentration (by weight) according to LC-MS is shown among samples that 1) did have lidocaine present on DART-MS, and with lidocaine quantified on LC-MS, 2) without xylazine present on DART-MS (meaning all samples should be negative on xylazine test strip), and 3) with xylazine test strip results. A log scale is used on the x-axis to show the percent concentration by mass of lidocaine. Results are shown separate by false positive (shown in red) and true negatives (shown in blue). This figure highlights that false positives, which have been shown in other studies to occur in the presence of lidocaine, occurred at a variety of lidocaine concentrations.

**Supplemental Table 3.** Sample Sizes of Combinations of Testing Methodologies Used For all samples considered in this study, each combination of testing methodologies is shown as a row, along with the corresponding sample size. An explanation of the circumstances leading to each combination of testing methods is also provided.

| **XTS** | **DART-MS** | **LC-MS** | **Sample Size** | **Explanation of Circumstances** |
| --- | --- | --- | --- | --- |
| 1 | 1 | 0 | 410 | These samples had xylazine test strips performed in the field, and swabs were sent for DART-MS. LC-MS was not done, either because the participant was unable or unwilling to provide a sufficient mass of sample, or because quantification vials had run out for the day of data collection due to a limited supply when piloting the lab quantification procedures. |
| 0 | 1 | 0 | 384 | These samples had DART-MS testing done. Xylazine test strips were not done, either because participants did not want them to be used, or because they did not believe their sample would contain fentanyl (which would trigger xylazine strip testing). LC-MS was not done, either because the participant was unable or unwilling to provide a sufficient mass of sample, or because quantification vials had run out for the day of data collection due to a limited supply when piloting the lab quantification procedures. |
| 0 | 1 | 1 | 328 | These samples were sent for DART-MS and LC-MS but did not have xylazine test strips performed. This occurred when patients did not wish to engage in test strips, but accepted other testing modalities. |
| 1 | 1 | 1 | 305 | These samples had all testing methodologies used. Sufficient sample was available for LC-MS testing, and participants accepted all 3 modalities. |
| 1 | 0 | 0 | 86 | These samples had xylazine test strips performed, but participants declined confirmatory testing with DART-MS or LC-MS, likely given those modalities do not provide results in real time. |
| 0 | 0 | 0 | 57 | These samples had none of the testing modalities. Instead, they likely obtained only FTIR testing, or only non-xylazine test strips. |
|  |  | **Total** | **1570** |  |

